# Quantitative Shape Analysis Supports the Use of Aortic Arch Stent Grafts in Complex Aortic Geometries

**DOI:** 10.64898/2026.10.25.26365919

**Authors:** Kelly C. Twohig, Joseph A. Pugar, Ian McDougall, Cheong Jun Lee, Ross Milner, Luka Pocivavsek

## Abstract

**Introduction:** Traditional thoracic endovascular aortic repair (TEVAR) relies on distal landing zones beyond the left common carotid artery, which may underperform in patients with complex aortic anatomy. The Nexus Aortic Arch Stent Graft enables proximal landing zones in the ascending aorta, expanding endovascular options for challenging arch pathology. This study applies a geometrically focused framework using fluctuation in total integrated Gaussian curvature (*δK*), a size-invariant metric of surface topology, to evaluate Nexus performance across diverse aortic morphologies.

**Methods:** Pre-operative CT imaging was analyzed for 27 patients treated with the Nexus device and 37 treated with traditional TEVAR (proximal landing zones 2/3). CT scans from 215 patients with non-pathologic aortas served as controls. Two-dimensional aortic surface meshes were constructed to calculate *δK* and mean aortic radius, while tortuosity index was derived from the centerline. Nexus data were mapped into a previously established size-shape feature space to assess predicted outcomes. TEVAR failure was defined as reintervention or death due to aneurysmal growth, endoleak, dissection extension, or rupture. Correlation and clustering analyses evaluated relationships between aortic morphology and outcomes. A scale-sampling approach captured the multiscale nature of aortic geometry while reducing sensitivity to mesh resolution and partitioning.

**Results:** The Traditional TEVAR cohort had a significantly lower success rate than the Nexus cohort (51.4% vs. 85.2%, p = 0.005). Traditional TEVAR was particularly less effective in patients with elevated *δK* and large mean radius, whereas the Nexus device demonstrated favorable outcomes even in anatomically complex aortas with similarly elevated *δK* and radius values. Combining *δK* and radius demonstrated distinct clustering patterns by pathology: 78.3% of chronic dissections occupied the high *δK*/radius zone, while 70.6% of isolated aneurysms fell in the low *δK*/radius zone (p <0.001). *δK* provided unique morphological insight beyond size, whereas tortuosity index was less discriminative.

**Conclusion:** δ*K* offers a quantitative framework for assessing device performance across variable aortic anatomy. Endovascular arch repair using proximal ascending aortic landing zones, exemplified by the Nexus device, may provide a viable option in patients with complex geometry. This shape-size framework represents a step toward personalized patient selection and improved long-term outcomes.

## Introduction

Management of aortic arch pathology remains a significant clinical challenge, although treatment options have expanded considerably in recent years (1,2). The approach to treating aortic arch disease, including aneurysm and dissection, falls into three categories: open surgical repair, endovascular repair, and hybrid repair. Open surgical total arch replacement remains the conventional method but requires cardiopulmonary bypass and hypothermic circulatory arrest (2). Hybrid approaches can circumvent these requirements but still involve median sternotomy, making them unsuitable for fragile patients with comorbidities or prior thoracic surgeries. Total endovascular arch repair has emerged as a less invasive approach, eliminating the need for cardiopulmonary bypass, circulatory arrest, or median sternotomy. According to guidelines from the American College of Cardiology and the American Heart Association, complete endovascular repair of aortic arch aneurysms may be reasonable for high-risk patients (3).

However, complete endovascular repair of the aortic arch presents secondary to anatomic constraints: proximity to the great vessels increases stroke risk, and arch curvature challenges the creation of adequate landing zones (4,5). Recent innovations in arch stent grafts aim to overcome these restrictions by introducing branches or fenestrations that restore blood flow to the brain and upper extremities (6–8). Prior to device implantation, extra-anatomical bypasses can be performed via supraclavicular incisions, mitigating the need for sternotomy. One promising addition to this cohort is the Nexus Aortic Arch Stent Graft System (Endospan, Herzilya, Israel) – a single-branch, two-stent graft system designed for zone 0/1 and intended for high-risk or fragile patients with aortic arch pathologies (9). Given their ability to completely seal and depressurize the aortic arch, stent grafts with proximal landing zones in the ascending aorta not only provide a novel and less invasive approach to treating arch disease but also have the potential to address aortas with significant shape deviations.

There is growing interest across the medical field in leveraging shape analysis to gain deeper insights into anatomical structures and disease progression. Geometry has been used to predict osteoarthritis risk based on hip structure, estimate survival probabilities from brain tumor morphology, and assess the extent of cardiac remodeling after myocardial infarction through statistical shape analysis of the left ventricle (10–12). There is a unique opportunity in vascular surgery to harness the vast amount of shape information obtained from cross-sectional imaging to drive innovation in device development and provide robust tools for evaluation.

More advanced statistical shape analysis (SSA) approaches have also been applied to aortic morphology, including atlas-based and principal component-based methods capable of capturing complex geometric variation across patient populations (13–16). These approaches provide high-dimensional characterization of anatomy but may be computationally intensive and less readily interpretable in clinical settings. In contrast, the present work focuses on a simplified and clinically interpretable size-shape framework based on mean radius and fluctuation in total integrated Gaussian curvature (*δK*).

While axial diameter remains the standard for evaluating aortic pathology, geometric analysis offers a more comprehensive view of the aorta’s heterogeneous, three-dimensional morphology. Metrics such as tortuosity index, question mark angle, and mean centerline curvature quantify aortic shape from the centerline (17–19). Because the diseased aorta most often deforms in a heterogeneous manner, shape metrics calculated from the centerline lose critical information about the true morphology of the aortic surface. More recently, fluctuation in total integrated Gaussian curvature (*δK*) has been introduced as a novel size-invariant measurement that quantifies aortic shape based on surface topology (20).

*δK* captures the degree to which the aorta deviates from uniform three-dimensional shapes like cylinders or spheres. Because the aorta is topologically equivalent to a bent cylinder, the sum of curvature *K* is constant. If a cylinder is deformed by creating a local outpouching, positive curvatures (convex areas) are balanced by negative curvatures (concave areas). Thus, when an aorta deforms, every increase in *K* is proportionally offset by a decrease elsewhere on the aortic surface. The variance in curvature (*δK*) captures both positive and negative curvatures, thus carrying detailed information about aortic shape. A visual depiction of both radius and *δK* is presented in **Figure 1**.

**Figure 1:**
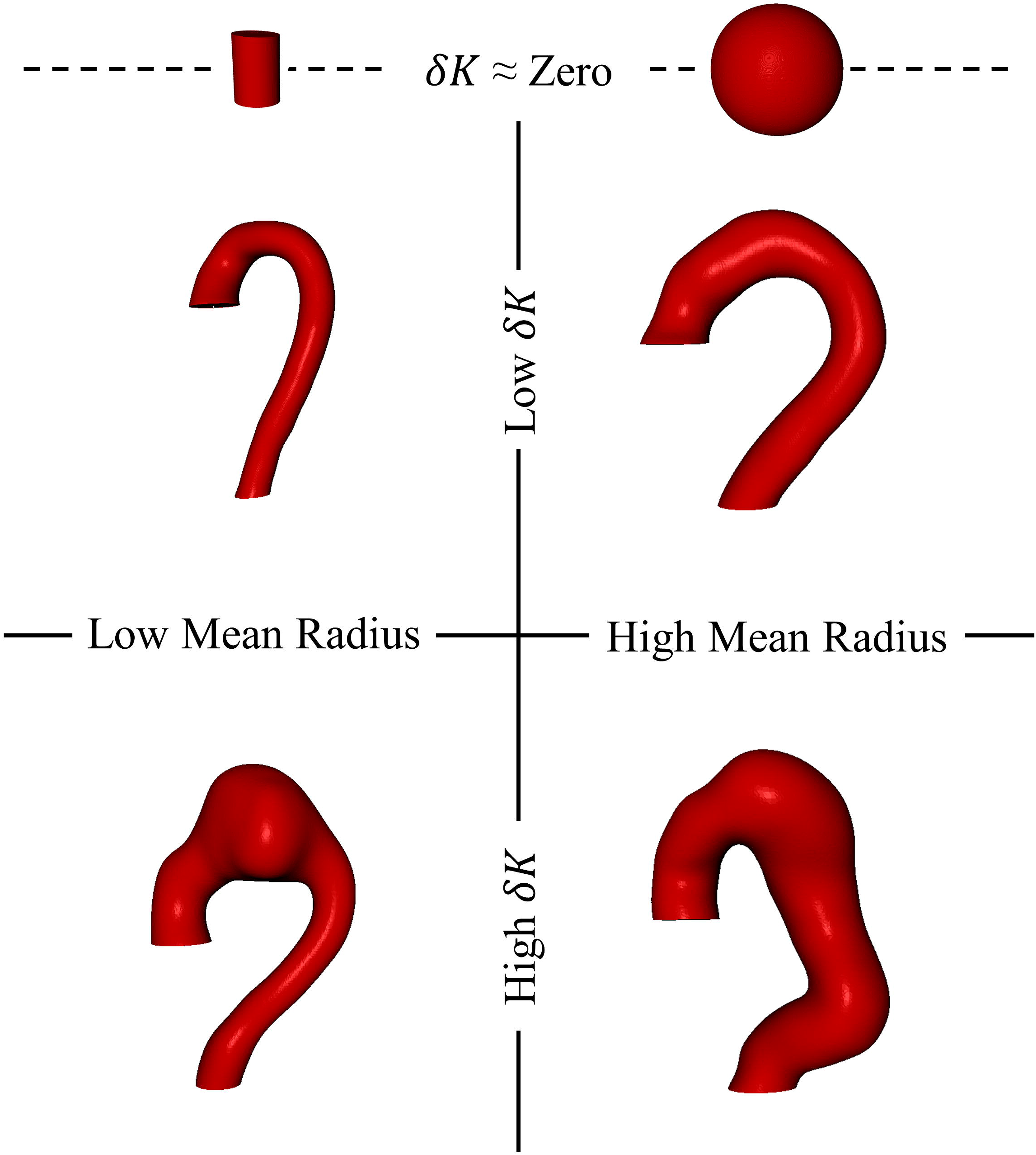
A visual depiction of fluctuation in total curvature (*δK*). *δK* captures variations in aortic shape by quantifying changes in curvature at points across its entire surface. A perfectly uniform structure, such as a sphere or a cylinder, will have a *δK* of zero. A smaller or larger uniform structure will change along the size axis but its *δK* value will remain constant (dotted line). A normal, non-pathologic aorta is topologically equivalent to a bent cylinder (upper left). If the normal aorta deforms or grows uniformly, it increases in mean aortic radius with minimal to no changes in *δK* (upper right). If the aorta deforms heterogeneously, as is commonly seen in aneurysmal degeneration, *δK* will increase. *δK* can quantify the extent of shape degeneration whether it occurs locally, such as in an isolated aortic arch aneurysm (bottom left), or globally, as in a fusiform thoracic aortic aneurysm (bottom right).

*δK* has shown improved performance in classifying aortas into non-pathologic/normal, successful thoracic endovascular aortic repair (TEVAR), and failed TEVAR categories compared to other shape measurements (20). Machine learning models incorporating both *δK* and aortic size have achieved classification accuracies of 97%, 71%, and 88% for these categorical assignments, respectively, outperforming models based on size or shape alone (21).

In this study, we apply *δK* as a novel way to quantitatively categorize aortic arch anatomy and assess the efficacy of aortic arch endovascular stent grafts in treating aortas with extreme shape deviations. We aim to evaluate whether this procedural approach provides a viable treatment option for aortas with a combination of high *δK* and high aortic diameter.

## Methods

### Clinical Cohort and Segmentation

The Nexus device was previously trialed through a prospective, multi-center study at three centers in Europe, which included 18 patients under a feasibility clinical study, and through an observational study at four centers in Europe, Canada, and New Zealand, which included 10 patients under compassionate use procedures (9,22). Pre-operative computed tomography (CT) scans were obtained from all 28 patients through a Data Use Agreement between Endospan Ltd. and the University of Chicago.

The single-institution cohort consists of 37 patients who underwent TEVAR by five surgeons at the University of Chicago and Endeavor Health Evanston Hospital. None of these patients had a history of connective tissue disease. All TEVAR patients had proximal landing zones in aortic zones 2 or 3, and only pre-operative CT scans were analyzed. The institutional cohort also included 215 patients with non-pathologic aortas who presented to the University of Chicago trauma surgery service and underwent CT angiography of the chest. Patients with traumatic aortic injury or other known aortic pathology were excluded from this cohort. These groups are referred to as the “Traditional TEVAR” and “Normal” cohorts throughout the analysis. All collection and analysis followed the Declaration of Helsinki guidelines and institutional review board approval (IRB20-0653, IRB21-0299). Imaging datasets consisted primarily of contrast-enhanced CT angiography studies, although contrast-enhanced CT scans were also included in select cases. Due to the retrospective multi-institutional nature of the dataset, acquisition parameters varied between scans; however, all studies included slice thicknesses <5 mm to permit adequate three-dimensional segmentation and geometric analysis.

Demographic and outcomes data were collected for all patients. Unsuccessful TEVAR was defined as the need for reintervention or death related to aneurysmal growth, endoleak, dissection extension, or rupture. Group comparisons were performed using Kruskal-Wallis, Mann-Whitney U, and chi-squared tests with statistical analyses conducted using SciPy in Python (23). Significance was defined as p ≤ 0.01.

Three-dimensional aortic models were generated from CT scans in Simpleware (Synopsys, Inc., Sunnyvale, USA), aided by a semi-automated aortic segmentation software (PRAEVAorta Web, Nurea, Bègles, France), which has undergone prior external validation in vascular imaging applications (24). Segmentations were bounded proximally by the right coronary artery and distally by the celiac artery. All segmentations underwent manual quality review prior to analysis, and scans with poor boundary definition or inadequate image quality were excluded. One Nexus patient CT scan was excluded due to poor resolution, which prevented adequate segmentation quality. Centerlines were automatically generated in Simpleware from the smoothed segmentation masks prior to tortuosity index calculation. Tortuosity index was calculated by dividing centerline length by the Euclidean distance between the centerline endpoints.

### Smoothing, Meshing, and Partitioning

The smoothing, meshing, and partitioning workflow used for *δK* analysis is illustrated in **Figure 2**. Raw segmentation masks were smoothed using a combination of mask dilation and recursive Gaussian filters. A two-dimensional surface mesh consisting of triangular elements was then generated for each segmentation mask. Each mesh was subsequently divided into a variable number of surface partitions proportional to aortic size. The mean radius (computed per partition using local inverse curvature values) and the total integrated Gaussian curvature (*K*) of each partition were calculated. For each aorta, the mean radius value and the fluctuation (variance) of K (*δK*) were derived from the distribution of surface patches. Normalized *δK*(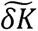), was calculated by dividing the *δK* of a given aorta by the average *δK* of the normal cohort.

**Figure 2:**
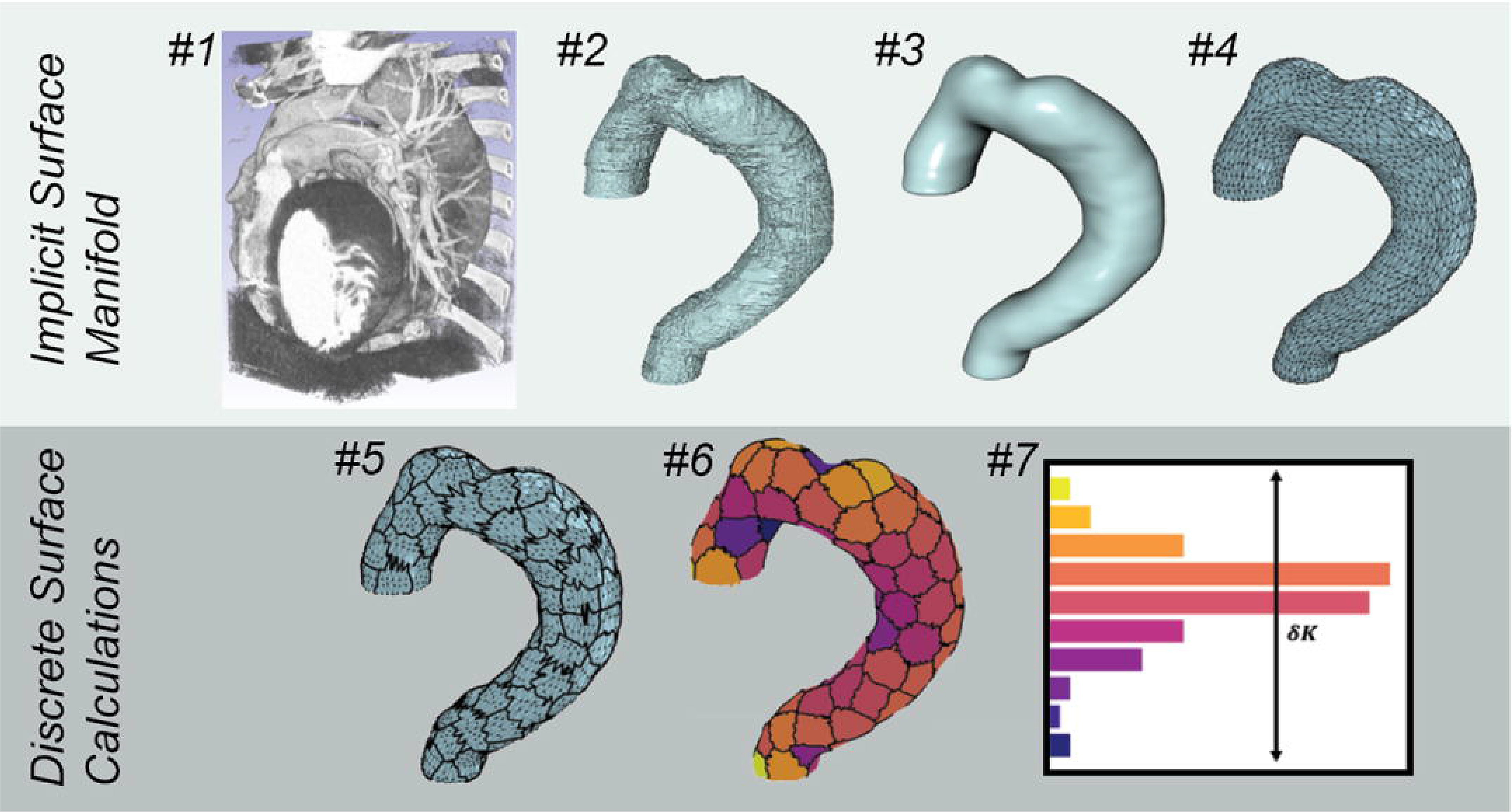
Representative workflow for segmentation, meshing, partitioning, and *δK* analysis of aortic geometry. (#1) Original CT angiographic DICOM reconstruction of the thoracic aorta. (#2) Initial aortic surface segmentation generated from CT imaging, demonstrating surface irregularity related to image voxel resolution. (#3) Smoothed aortic surface following application of recursive Gaussian surface smoothing (Gaussian length scale = 7 voxels). (#4) Triangular surface mesh generated from the smoothed segmentation mask (5000 triangular elements shown). (#5) Partitioned aortic surface following subdivision of the mesh into discrete surface patches using a K-means clustering algorithm (115 patches shown). Dark borders delineate individual surface partitions. (#6) Total integrated Gaussian curvature (*K*) mapped across the partitioned surface. Patch color corresponds to local *K* values, with lighter colors representing regions of higher integrated curvature and darker colors representing regions of lower integrated curvature. (#7) Distribution of per-patch *K* values used to calculate fluctuation in total integrated Gaussian curvature (*δK*). Segmentation and mesh generation were performed in Simpleware ScanIP, while partitioning and *δK* calculations were performed using Python-based analysis tools.

Each smoothed aortic mask was meshed and partitioned using six different discretization length scales to create six unique 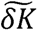 values per patient scan (21). This scale-sampling approach was used to better capture the multiscale nature of aortic geometry while reducing sensitivity to mesh resolution and partitioning choice. The number of surface partitions was scaled proportionally to aortic size to preserve comparable spatial sampling across differently sized aortas. The differing scales used 5 mm² and 10 mm² element meshes combined with partition pre-factors of 0.5, 1, or 5, resulting in partitioning schemes averaging approximately 50, 100, and 500 surface patches. These specific combinations were previously validated to maximize signal preservation and interpretability while minimizing discretization-related noise and computational burden (21).

### Correlation and Clustering Analysis

Rank-order correlation analysis assessed relationships between variables, specifically 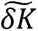, mean aortic radius, and tortuosity index. Pearson’s correlation coefficient evaluated the strength of linear relationships, while Spearman’s rank correlation coefficient quantified monotonic relationships. Additionally, clustering analysis was performed on the increasing 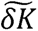 and mean radius series to evaluate data grouping. This analysis aimed to determine whether clear separation or trends existed in aortic morphologies across the dataset, using clustering algorithms that optimize group distinctiveness and minimize intra-cluster variation.

### Feature Space Classification

Data points from the Normal and Traditional TEVAR cohorts were plotted into a two-dimensional feature space, with aortic size (mean aortic radius) on the x-axis and aortic shape (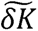) on the y-axis. A multinomial logistic regression model was trained and tested using 20 unique test splits (80% training / 20% testing). Aortic size and shape were used as independent input features, while aortic classification (normal, successful TEVAR, and failed TEVAR) was the dependent variable. The F1-score of the model, computed as the harmonic mean of precision and recall, was used to measure accuracy given class disproportionalities.

This three-class predictive machine learning model has been described in previous work (16, 17), but the institutional data set was modified for this analysis to include only patients with TEVARs deployed distal to zone 1. After re-establishing the three background boundary zones, Nexus patient data points were plotted into the feature space to evaluate aortic morphology and correlate with TEVAR outcomes.

## Results

Demographic data for the three cohorts are outlined in **Table 1**. Nexus patients were, on average, older than the Traditional TEVAR and Normal cohorts (72.3 years ± 6.2 years vs. 61.2 years ± 15.2 years vs. 53.2 years ± 20.1 years, p < 0.001). Nexus patients were also significantly more likely than Traditional TEVAR patients to have undergone a previous sternotomy (55.6% vs. 21.6%, p = 0.005). Procedure information is detailed in **Table 2**.

**Table 1:**
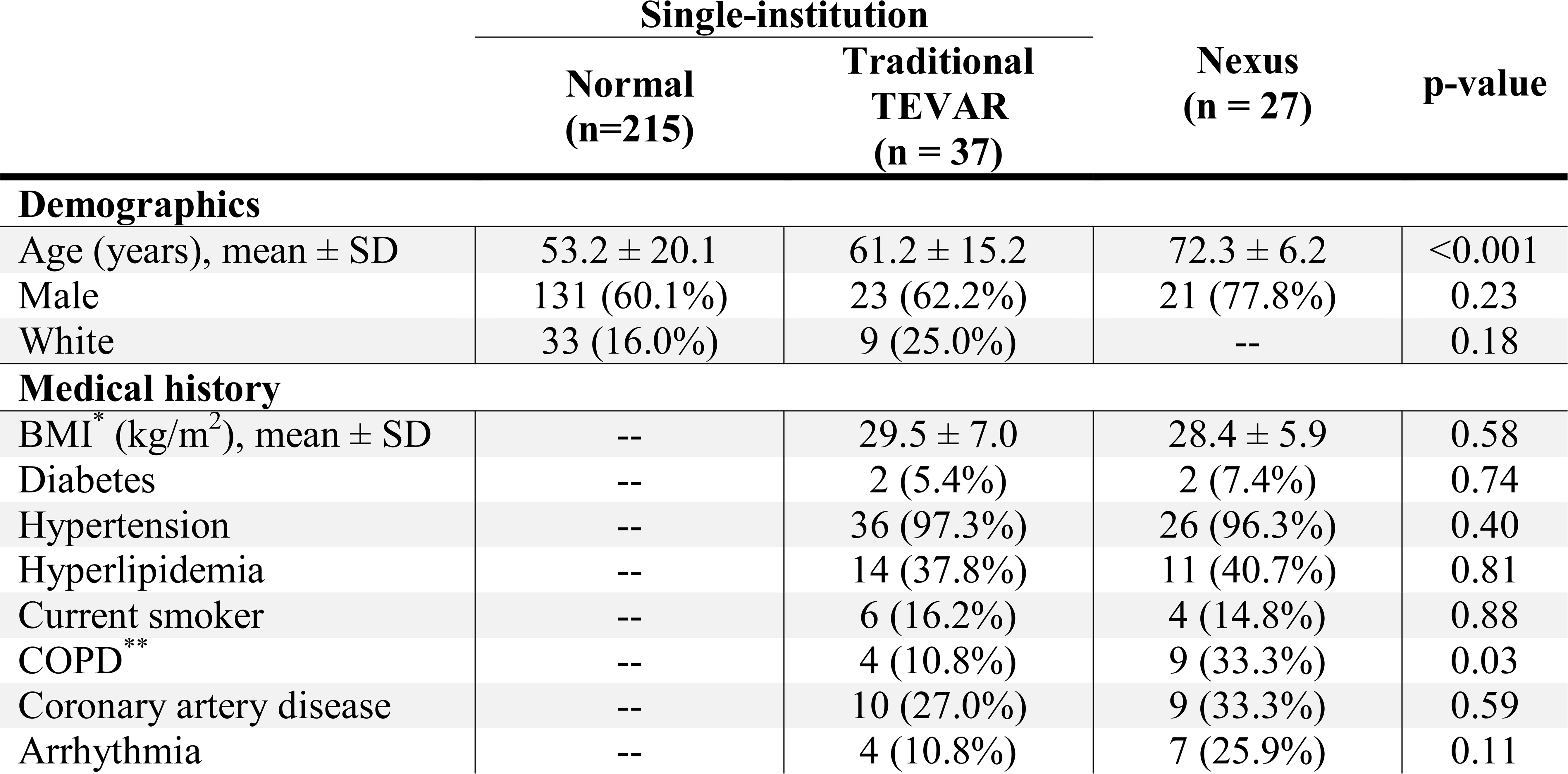
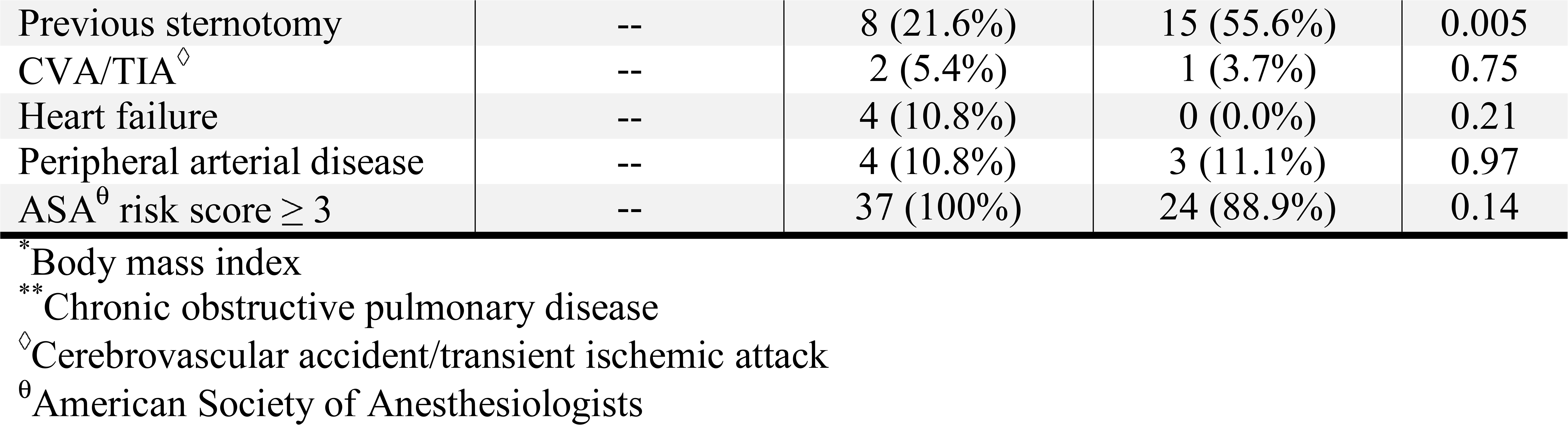
Demographic and comorbidity data of 252 patients from an institutional cohort (215 with non-pathologic aortas, 37 with pathologic aortas requiring TEVAR) and 27 patients from the Nexus European Union trial. All Traditional TEVAR patients had proximal landing zones in aortic zones 2 or 3, and all Nexus patients had proximal landing zones in aortic zone 0. Complete comorbidity data were not consistently available for the trauma-derived normal cohort. All data points are represented as n (%) unless otherwise stated.

**Table 2:**
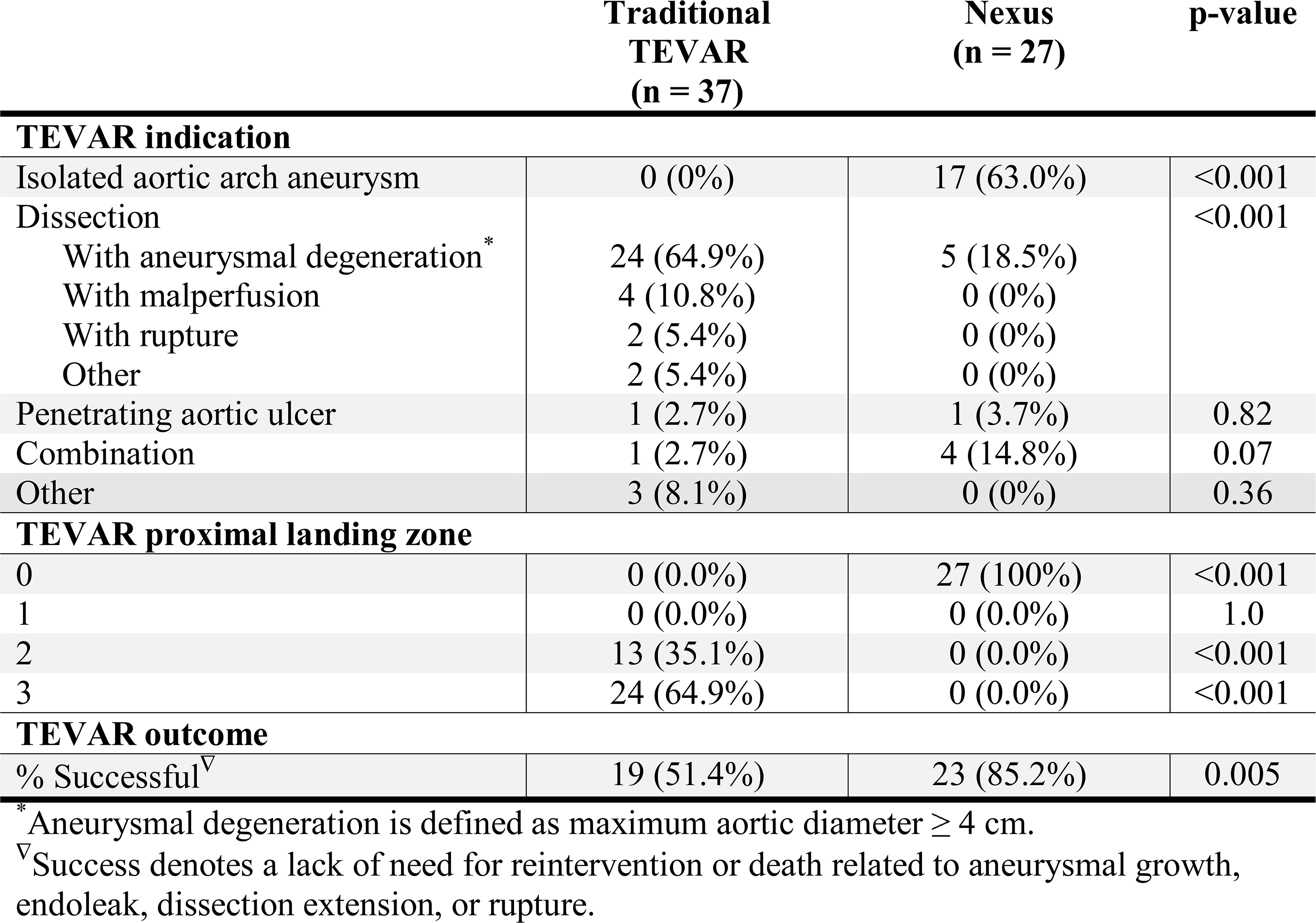
Procedure information for 37 patients that underwent traditional TEVAR and 27 patients that underwent TEVAR through the Nexus European Union trial. All data points are represented as n (%).

Nexus patients most commonly underwent TEVAR for an isolated aortic arch aneurysm (63.0%), while the Traditional TEVAR cohort most frequently presented with chronic aortic dissections resulting in aneurysmal degeneration (64.9%). All Nexus patients had proximal TEVAR landing zones in zone 0 (ascending aorta), whereas all Traditional TEVAR patients had proximal landing zones in zone 2 (distal to the left common carotid artery) or zone 3 (distal to the left subclavian artery). About half (51.4%) of Traditional TEVAR patients had successful outcomes without the need for reintervention or aortic-related death secondary to aneurysmal growth, endoleak, dissection extension, or rupture. The Traditional TEVAR cohort demonstrated a lower rate of freedom from reintervention or aortic-related death compared to the Nexus cohort (85.2%), although these groups differed substantially in underlying pathology and procedural indication.

The 27 pre-operative aortas of the Nexus patients are displayed in **Figure 3** and **Figure 4**. These models were generated utilizing the segmentation and smoothing processes detailed above. Aortas associated with unsuccessful TEVAR outcomes are highlighted with numbered circles. **Figure 3** arranges the aortas by increasing 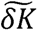. 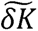 is a global topological parameter that evaluates curvature changes across the entire aortic surface, treating isolated saccular aneurysms and larger fusiform aneurysms equivalently. Qualitative inspection demonstrated noticeable differences in aortic shape as 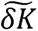 increases. Aortas with relatively low 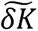 values (blue) predominantly exhibit focal, saccular arch aneurysms. As 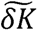 values increase (green/yellow), the aortas demonstrate greater tortuosity, with larger or more numerous saccular aneurysms. At the highest 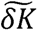 values (orange/red), the aortas demonstrated pronounced tortuosity, multiple saccular aneurysms, and/or extensive fusiform dilation.

**Figure 3:**
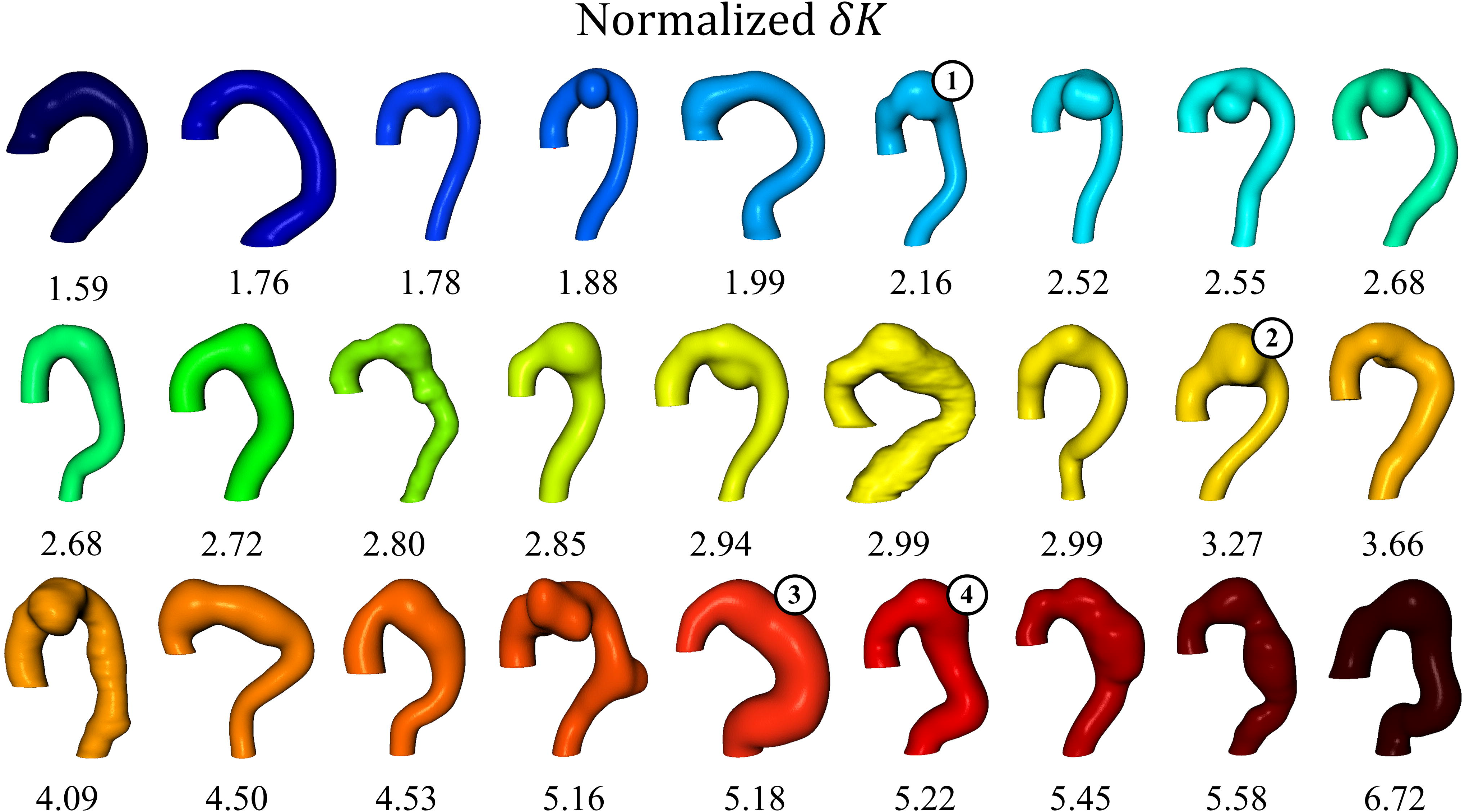
Three-dimensional models of the pre-operative aortas of 27 Nexus patients organized from left to right by increasing normalized *δK*(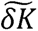), generated from CT imaging. These models were created by smoothing segmentation masks with a combination of dilation and recursive Gaussian filtering. A numbered circle indicates that the patient had an unsuccessful TEVAR. The color mapping corresponds to 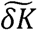 values, with blue representing low 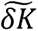 and red indicating high 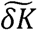. The 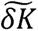 value is displayed below each aorta for reference.

**Figure 4:**
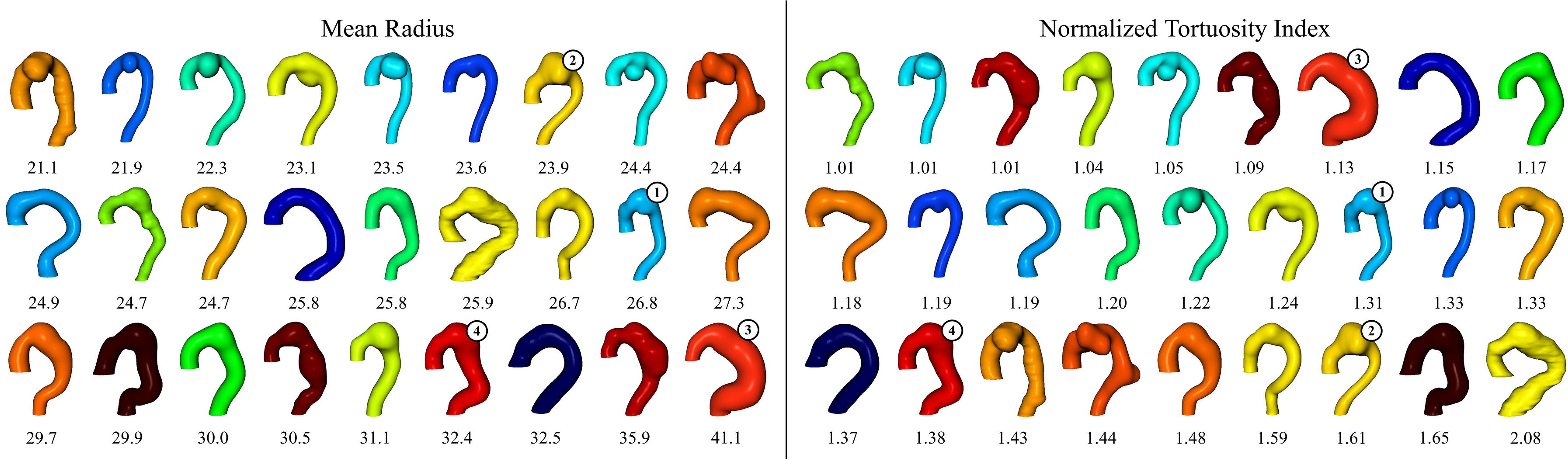
The left panel arranges the same aortas from Figure 3 by mean radius, and the right panel sorts by normalized tortuosity index. Again, a numbered circle indicates unsuccessful TEVAR. Color mapping from Figure 3 is retained, corresponding to each aorta’s 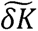. The mean radius and normalized tortuosity index are displayed below each aorta, respectively.

The left panel of **Figure 4** organizes the same aortas by increasing mean aortic radius, and the right panel of **Figure 4** sorts by normalized tortuosity index (right). Increasing mean aortic radius organizes the aortas sequentially, with smaller, locally dilated aortas at the front of the sequence and more uniformly dilated aortas toward the end. Notably, aortas with multiple focal saccular aneurysms or larger saccular aneurysms are more broadly distributed throughout the group, resulting in a sequence that differs from **Figure 3**. Conversely, visual inspection suggests that ordering by tortuosity index provides limited morphologic discrimination, with aortas of varying shapes and sizes appearing intermixed. To quantitatively evaluate these ordering relationships, rank-order correlation analysis was performed between 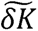, mean radius, and tortuosity index.

A rank-order correlation revealed moderate positive monotonic and linear relationships between 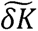 and mean radius, with a Pearson correlation coefficient of 0.48 and a Spearman correlation coefficient of 0.39. In contrast, weak linear and monotonic relationships were observed between the tortuosity index and mean radius (Pearson: -0.14, Spearman: -0.13). These findings indicate that 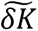 and mean radius provide complementary information about the morphology and size of the aorta, but because both correlation coefficients are well below one, the relationship is moderate at best. This suggests that aortic shape (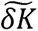) provides information that is at least partially independent of size (mean radius). Cluster analysis of the sequences from **Figure 3** and **Figure 4** showed distinct clusters with clear separation in the 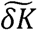 series, continuous scaling in the size series, and minimal grouping in the tortuosity index series. These findings quantitatively demonstrate that tortuosity index exhibits weak relationships with other measures of aortic morphology and provides limited discriminatory capability for differentiating between aortic anatomies.

Information about pre-operative aortic shape and size is summarized in **Tables 3a** and **3b**. There was no significant difference in average mean aortic radius between the two TEVAR groups (26.0 mm ± 4.0 mm vs. 27.2 mm ± 4.6 mm, p = 0.52). Although the Nexus group trended toward a higher mean 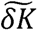, the difference was not statistically significant (2.8 ± 1.0 vs. 3.4 ± 1.4, p = 0.06). Likewise, within the Nexus cohort, the chronic dissection patients trended toward a higher mean 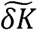 compared to the isolated arch aneurysm/penetrating aortic ulcer (PAU) patients, however the difference was not statistically significant (3.8 ± 1.7 vs. 3.2 ± 1.2, p = 0.47).

**Table 3a:**
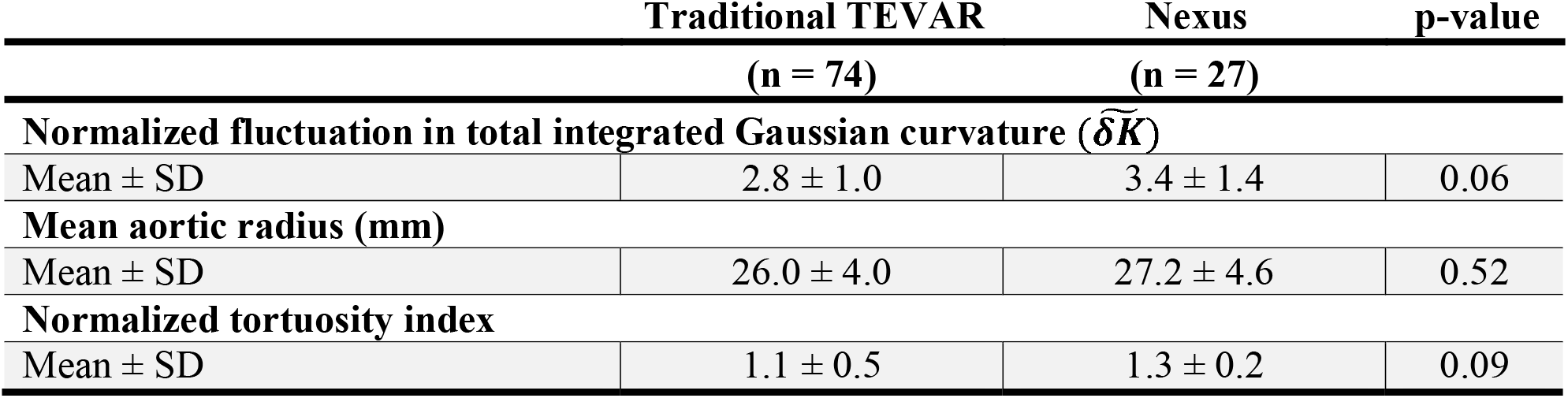
Aortic shape (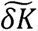), aortic size (mean aortic radius), and normalized tortuosity index extracted from pre-operative CT imaging. Many patients in the Traditional TEVAR cohort underwent multiple CT scans; therefore, this group includes 74 pre-operative CT scans, with each scan represented once. These values were calculated from meshes with a density of 10 mm^2^ and a partition pre-factor of 1. For reference, the mean 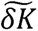 of the normal population (n=329) was 1.0 ± 0.3, the average mean aortic radius was 18.1 mm ± 2.9 mm, and the mean normalized tortuosity index was 1.0 ± 0.2.

**Table 3b:**
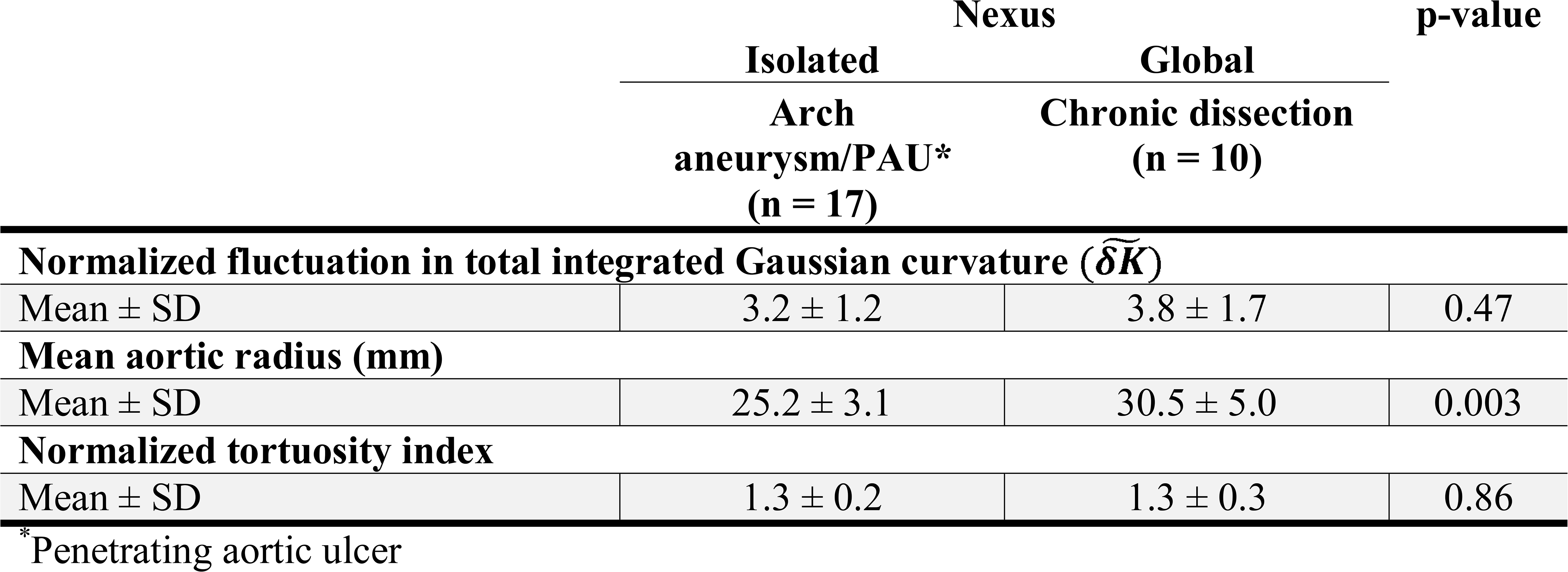
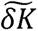, mean aortic radius, and normalized tortuosity index for the Nexus cohort, now stratified by pathology. Isolated pathologies include arch aneurysms and penetrating aortic ulcers, while global pathologies include chronic dissections with or without aneurysmal degeneration.

**Figure 5** illustrates the normal and pathologic aortas from the single-institution cohort, plotted in a two-dimensional feature space defined by aortic size (mean aortic radius) and shape (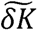). A multinomial logistic regression model trained on this data achieved an F1-score of 0.91. The learned decision boundaries are depicted in the background. Nexus cohort data points were overlaid onto these boundaries (**Figure 6**), with a total of 162 data points plotted (six per pre-operative CT scan from the scale-sampling methodology (17)). Panel A shows all data points, color-coded according to their corresponding aortic models from **Figures 3 and 4**. Panel B highlights the failed Nexus TEVAR points. In total, 85 points (52.5%) fell within the feature-space region associated with successful Traditional TEVAR outcomes (blue), and 79 of these (92.9%) corresponded to successful Nexus outcomes. The remaining 77 points (47.5%) fell within the feature-space region associated with failed Traditional TEVAR outcomes (red); however, only 18 (23.3%) corresponded to failed Nexus outcomes, while 59 (76.6%) remained successful. Panel C stratifies the Nexus patients by pathology: purple points represent globally pathologic aortas (e.g., chronic dissections), while yellow points represent isolated pathologies (e.g., isolated aortic arch aneurysms and PAUs). Globally pathologic aortas more frequently occupied the feature-space region associated with failed Traditional TEVAR outcomes, with 47 points (78.3%) located in the red region, whereas isolated pathologies more commonly occupied the region associated with successful Traditional TEVAR outcomes, with 72 points (70.6%) located in the blue region (p < 0.001).

**Figure 5:**
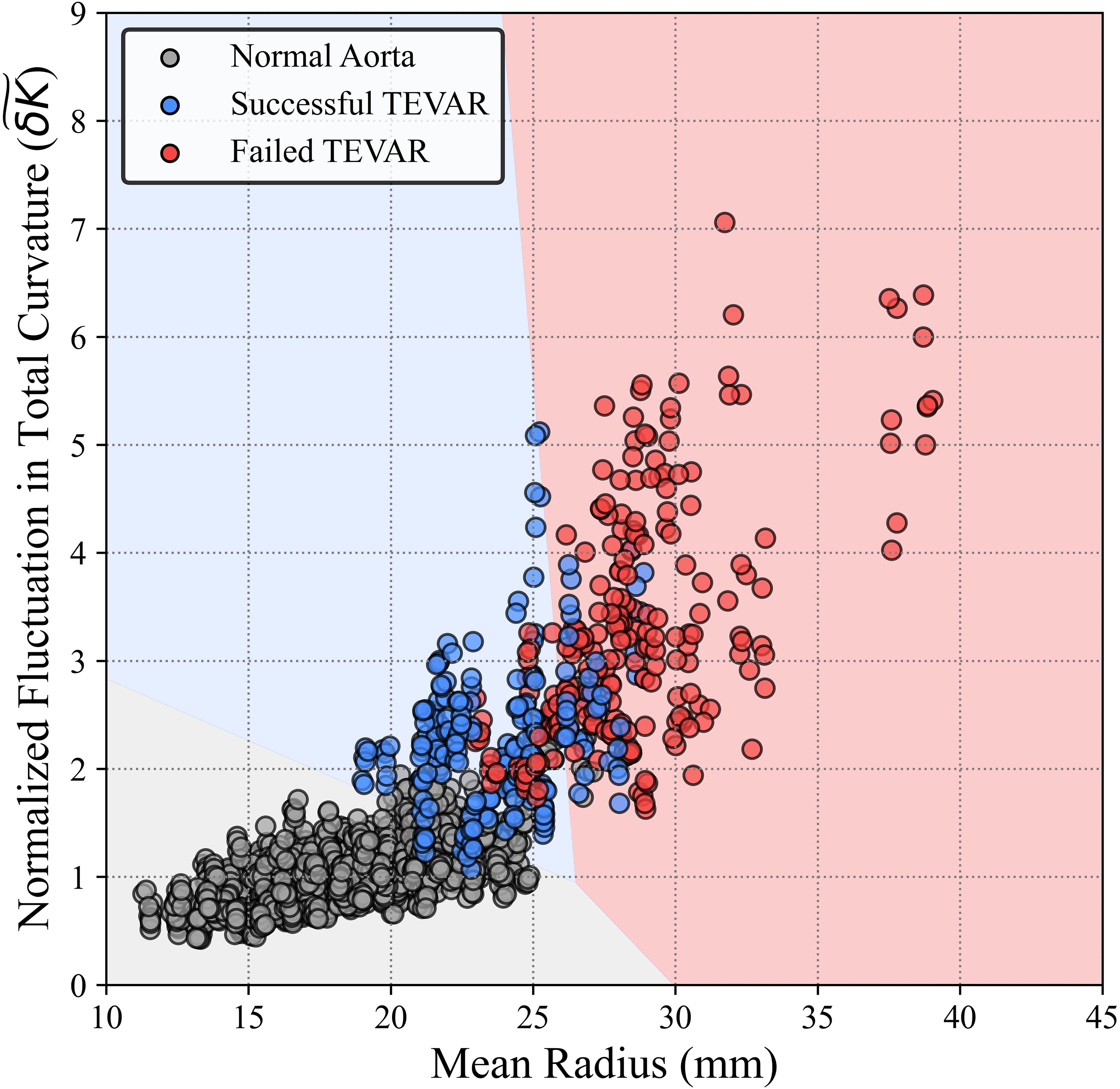
The Normal and Traditional TEVAR cohorts plotted into a two-dimensional feature space with aortic size (mean radius) and shape (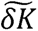). Each circle represents one aorta. Each aorta was meshed and partitioned in six different ways (mesh density = 5 or 10, partition pre-factor = 0.5, 1, or 5) to capture a full range of signal length scales. The background learned decision boundaries were created using a multinomial logistic regression model with F1-score = 0.91.

**Figure 6:**
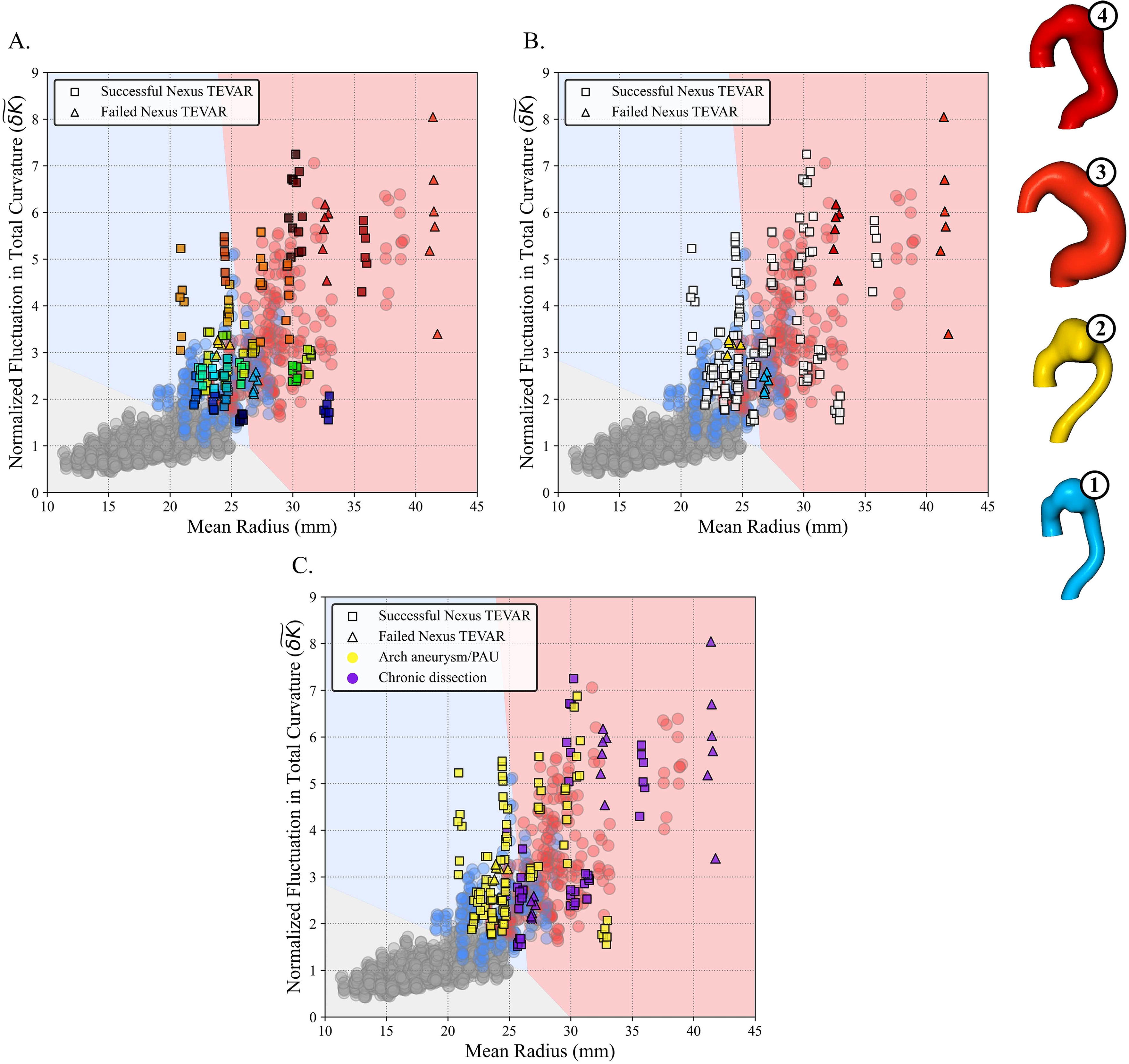
The Nexus dataset is plotted onto the decision boundaries established in Figure 5. As in Figure 5, each aorta was meshed with triangular mesh densities of 5 mm² and 10 mm² and partitioned using pre-factors of 0.5, 1, and 5. The blue background region represents the feature-space region associated with successful Traditional TEVAR outcomes, while the red background region represents the feature-space region associated with failed Traditional TEVAR outcomes. Successful Nexus TEVARs are represented by squares, while failed TEVARs are indicated by triangles. In Panel A, each Nexus patient is represented by a unique color corresponding to the color-coded aortic models shown in Figures 3 and 4 and ordered by increasing 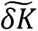. In Panel B, successful Nexus TEVARs are shown in white to emphasize the distribution of failed Nexus TEVARs. Additionally, each Nexus aorta with an unsuccessful outcome is displayed on the far right, with colors matching the corresponding plotted points. In Panel C, aortas are stratified by pathology: purple points represent globally pathologic aortas (e.g., chronic dissections), while yellow points represent isolated pathologies (e.g., isolated aortic arch aneurysms and PAUs).

## Discussion

This analysis presents a unique way to apply pre-operative aortic shape to understand how a novel device may fare in patients with a wide range of aortic geometries. For patients with increased mean radius and elevated 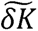, traditional TEVAR with zone 2 or 3 landing zones commonly resulted in the need for reintervention or death secondary to aortic disease. However, these findings suggest that TEVAR grafts that allow for a proximal landing zone in the ascending aorta, such as the Nexus stent graft, may be effective in patients with marked shape deviation (high 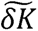) and increased size (high mean radius).

The specific outcomes of each Nexus patient with an unsuccessful TEVAR are detailed in **Table 4**. Aorta 2 required a TEVAR extension between 1 and 3 years post-procedure due to stent graft migration and development of a type IB endoleak. This was the only Nexus patient with an unsuccessful outcome whose data points primarily occupied the feature-space region associated with successful Traditional TEVAR outcomes, suggesting that this patient’s aortic morphology more closely resembled anatomies that historically demonstrated favorable outcomes with conventional TEVAR approaches.

**Table 4:**
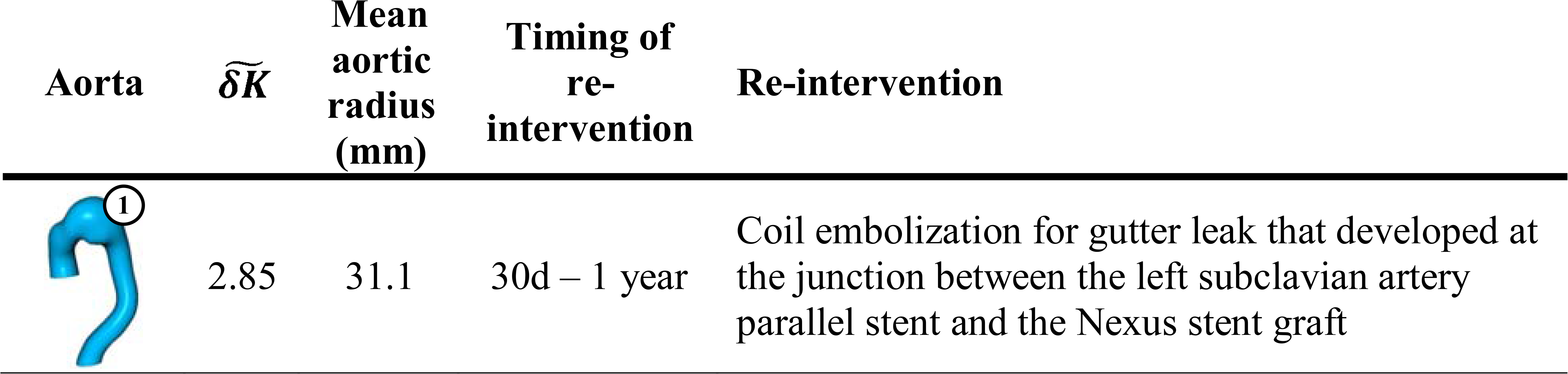
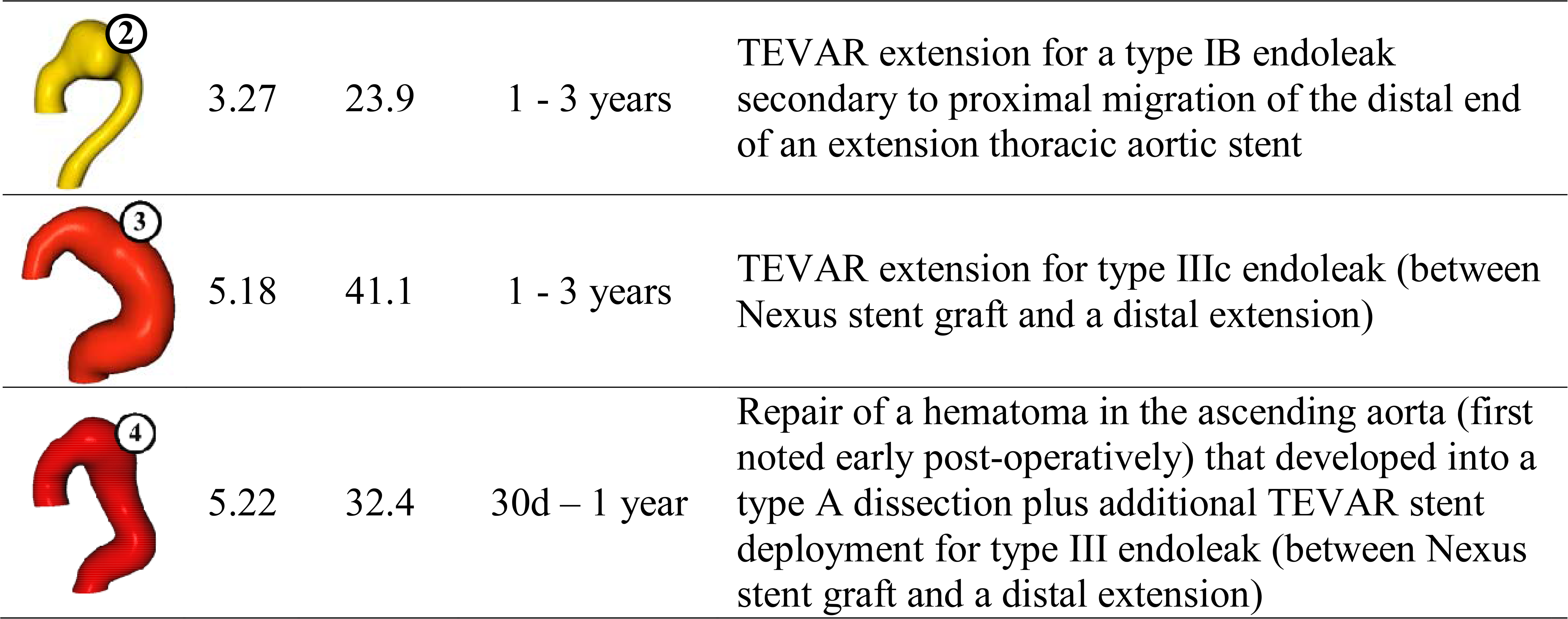
Each Nexus aorta with an unsuccessful outcome listed with its, mean aortic radius, and timing/description of re-intervention.

Of the three patients whose data points primarily occupied the feature-space region associated with failed Traditional TEVAR outcomes, one patient (Aorta 1) required coil embolization 4 months post-TEVAR for a gutter leak at the junction between the aortic stent graft and the left subclavian artery periscope stent. Between 30 days and 1 year post-procedure, Aorta 4 underwent repair of an ascending aortic hematoma that progressed to a type A dissection and required additional TEVAR stent deployment for a type IIIc endoleak between the Nexus stent graft and a Gore distal extension. Finally, between 1 and 3 years post-procedure, Aorta 3 required TEVAR extension for a type IIIc endoleak occurring between the Nexus stent graft and a Gore distal extension.

It is important to note that TEVAR indications differed between the two procedural groups. Nexus patients were more likely to undergo TEVAR for isolated aortic arch aneurysm, while the Traditional TEVAR patients most commonly underwent TEVAR for chronic dissection with an entry tear in the distal aorta. Notably, when TEVAR was indicated for chronic dissection, the entry tear was covered by the stent graft in all patients. Because of these differences in pathology and procedural strategy, comparisons in freedom from reintervention and aortic-related death should be interpreted cautiously, as they may reflect differences in underlying disease and treatment indication in addition to procedural approach. Accordingly, this comparison should be interpreted as an observational evaluation of morphology within two clinically distinct treatment paradigms rather than as a direct comparison of device efficacy. Nevertheless, the observation that many chronic dissection patients undergoing conventional TEVAR required subsequent reintervention raises the broader question of whether coverage of the intimal tear alone is sufficient in anatomically complex aortas.

A 2-cm proximal seal zone of healthy aortic tissue is widely recommended for TEVAR. This may be difficult to achieve in type B aortic dissection (TBAD) regardless of the location of the intimal tear, as intramural hematoma can often extend into the aortic arch. In a retrospective review of 71 patients who underwent TEVAR for TBAD at a single institution, only six (8.5%) achieved a 2-cm proximal seal zone free of intramural hematoma (25). Subsequent analysis revealed that 43.7% of patients would have required coverage of all three great vessels to achieve an adequate 2-cm landing zone of fully healthy aortic tissue (25). The concept of more proximal extension of the TEVAR stent graft leading to improved long-term outcomes has also been explored. A retrospective review of 83 patients that underwent TEVAR for acute TBAD found that the need for aortic reintervention and freedom from aortic-related adverse events at 36 months were significantly lower in patients that underwent TEVAR with zone 2 deployment compared to zone 3 deployment (26). These findings provide some context as to why 48.6% of patients in the Traditional TEVAR cohort ultimately required reintervention or experienced aortic-related death. In contrast, 70% (7/10) of Nexus patients with chronic dissections achieved successful TEVAR outcomes at three years, suggesting a potential association between more proximal seal zones and favorable long-term outcomes.

Our size-shape (mean radius, 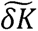)-feature space provides a quantitative framework for characterizing aortic morphology and contextualizing the extent of aortic arch coverage in patients without isolated arch pathologies. As shown in **Table 3b** and **Figure 6**, Nexus trial patients with chronic dissections exhibited elevated average 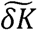 (3.8 ± 1.7) and mean radius (30.5 mm ± 5.0 mm), making comparison with the Traditional TEVAR cohort informative. In the Traditional TEVAR cohort, patients with similarly elevated 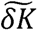 and mean radius frequently experienced aorta-related complications (20,21), corresponding to the feature-space region associated with failed Traditional TEVAR outcomes. However, 61.7% (29/47) of the Nexus chronic dissection data points occupying this same region corresponded to successful TEVAR outcomes.

These findings suggest that combining 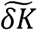 and mean radius captures important differences between focal aneurysms and more diffusely distributed aortic disease. More broadly, this framework identifies anatomical regions associated with increasing Traditional TEVAR failure rates while highlighting the potential utility of arch-specific devices in anatomically complex aortas.

This analysis specifically focused on aortic-related complications and outcomes associated with TEVAR. Of course, coverage of all branch vessels is not without its complications – of particular concern is the risk for stroke, along with other anticipated TEVAR complications such as renal failure and paraplegia. Endospan reported relatively low risks of these feared complications. Among the 28 patients studied, one (3.6%) experienced disabling stroke, one (3.6%) developed new onset renal failure requiring dialysis (although their renal function returned to baseline in two weeks), and zero suffered paraplegia (22). Another risk associated with coverage of the proximal aortic arch involves complications related to vessel debranching. Two patients required open surgeries for cervical bypass graft infections, and two others had asymptomatic cervical bypass occlusions without need for intervention (22). Additionally, although there were no deaths related to the Nexus device or aneurysm growth or rupture, Endospan reported an all-cause mortality rate of 27.6% at three years (22).

A limitation of this study includes the somewhat subjective nature by which TEVAR success is defined. Specifically, the need for intervention is determined by the individual surgeon and patient preferences. While aortic-related mortality serves as an objective endpoint, reintervention may vary between institutions and clinicians. Another limitation of this study is limited long-term data for the Nexus cohort beyond three years, preventing assessment of long-term durability. Additionally, the number of chronic dissection patients treated with the Nexus device was relatively small, limiting the precision of subgroup analyses. Future studies with extended follow-up and larger patient cohorts will be needed to validate these findings and further define the role of quantitative shape analysis in patient selection for advanced endovascular therapies.

## Conclusion

The Nexus Aortic Arch Stent Graft, which enables proximal landing in the ascending aorta, may represent a promising endovascular strategy for selected patients with highly complex aortic morphology. This study emphasizes the importance of quantifying aortic anatomy beyond simple size metrics, consistent with prior findings that centerline-based measures, such as tortuosity index, poorly differentiate aortic shapes (20). Given that aortic trials are often small and real-world aortic patients present with far more diverse anatomy, our work represents an important step toward enriching the anatomic characterization of aortic pathology.

Rather than replacing conventional CTA assessment, normalized 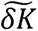 may serve as a complementary quantitative descriptor of aortic morphology during preoperative planning. For example, two patients with similar maximum aortic diameters may exhibit markedly different normalized 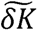 values, reflecting distinct anatomical phenotypes that are not captured by size alone and that may ultimately contribute to morphology-guided patient selection. Future prospective studies are needed to determine how quantitative shape metrics can best be incorporated into routine clinical decision-making.

With the rapid rise in the use of highly complex, data-driven machine learning approaches for patient selection and characterization, our size-shape features incorporate validated and clinically relevant domain knowledge into what are often very sparse dataset problems.

## Data Availability

All data produced in the present study are available upon reasonable request to the authors

## Notes

### Competing Interest Statement

Luka Pocivavsek reports financial support was provided by National Heart Lung and Blood Institute. Ian McDougall reports a relationship with Endospan, Ltd. that includes: employment and equity or stocks. Ross Milner reports a relationship with Endospan, Ltd. that includes: consulting or advisory. Endospan, Ltd., provided the data for analysis and helped in providing context of our findings within their prior published trial results. There was no direct financial support for this project from Endospan.

### Author Declarations

IRB of University of Chicago gave ethical approval for this work

### Funding

- National Heart Lung and Blood Institute: R01HL159205

